# The impact of the COVID-19 school closure on adolescents’ use of mental healthcare services in Sweden

**DOI:** 10.1101/2021.12.12.21267684

**Authors:** Helena Svaleryd, Evelina Björkegren, Jonas Vlachos

## Abstract

**Background:** School closures used to contain the COVID-19 pandemic may have negative impacts on students’ mental health but credible evidence is scarce. Sweden moved upper-secondary students to remote learning but, as the only country in the OECD, kept schools at lower levers open throughout the pandemic.

**Methods:** Using nationwide register data, we estimate the difference in the use of mental healthcare services between upper- and lower-secondary students during the pandemic, and relate this to the same group difference prior to the pandemic. For the main analysis, this difference-in-difference approach is applied to the period April-June 2020 when upper-secondary schools were fully closed. We also study the periods July-December 2020 when upper-secondary schools were largely open, and January-March 2021 when they were partially closed. We study the impact on all contacts with hospitals and specialist psychiatric care due to mental and behavioral disorder, as well as prescriptions for antidepressants, insomnia, and ADHD drugs.

**Findings:** Compared with expected rates, the use of mental healthcare services among upper-secondary students fell by -3.71 [CI95 -5.52 to -1.91] cases per 1000 during April-June 2020, largely due to a reduction in depression and anxiety-related diagnoses (-1.49; CI95 [-2.36 to -0.63]) and prescriptions (-1.80; CI95 [-2.93 to -0.68]). This reduction in the use of mental healthcare services corresponds to a 4.36% CI95 [-6.47 to -2.25]) decrease compared to the level prior to the pandemic. The decrease compared to expected rates persists through July-December 2020 (-3.55%; CI95 [-5.38 to -1.71]) and January-March 2021 (-5.23%; CI95 [-7.24 to -3.21]). The reduction is stronger among students in the 2^nd^ (-5.06%; CI95 [-8.02 to -2.09]) and 3^rd^ (-4.86%; CI95 [-8.19 to -1.53]) year of upper-secondary school. The decrease is concentrated to students who was not in contact with mental healthcare services earlier in the academic year (-16.70%; CI95 [-22.20 to -11.20]). The relative reduction is largest for unplanned care (-13.88%; CI95 [-19.35 to -8.42]) and care at emergency units (-18.19%; CI95 [-26.44 to -9.92]).

**Interpretation:** Closing upper-secondary schools in Sweden reduced use of mental healthcare services. There is no indication of this being due to reduced accessibility. In a setting with no strict lockdown, moving to online teaching for a limited period did not worsen mental health among students in upper-secondary schools.

## Introduction

School closures have been one of the most widely used disease containment measures during the COVID-19 pandemic. As education for billions of children has been disrupted there are raising concerns about the consequences of school closures on learning and mental health of children and adolescents (UNESCO 2021). Isolating the consequences of school closures on mental health is challenging as most countries closed all schools at the onset of the pandemic and at the same time introduced a number other types of non-pharmaceutical interventions (NPIs). In contrast to all other OECD countries, Sweden only closed upper-secondary schools for in-person schooling, while schools for younger students remained open (OECD 2021a). This partial school closure allows for a comparison of adolescents differently exposed to school closures but otherwise facing similar conditions. Using rich Swedish population wide register data, we study differences in the use of mental healthcare services between upper- (age 17-19) and lower- (age 14-16) secondary students during the pandemic. By comparing this to differences between the same groups prior to the pandemic, we adjust for long-term differences between upper- and secondary students and trends in use. Under the assumption that the use of mental healthcare services would have developed similarly for the two groups if upper-secondary schools had not closed, the estimated differences can be interpreted as causal.

This is the first study isolating the impact of school closures on use of mental healthcare services. There are, however, several studies from different countries of the development of healthcare use during the pandemic. For the initial phase of the pandemic, the evidence shows large decreases in the use of healthcare for self-harm, anxiety, depression, and for prescriptions of psychotropic drugs among adolescents (e.g. Jollant et al. 2021; Carr et al. 2021; Evensen et al. 2021; Ougrin 2020). After the initial phase, healthcare use seems to have reverted back to earlier levels in England (Carr et al. 2021) or possibly even higher levels in Norway (Evensen et al. 2021). A similar pattern is found for suicide in Japan (Tanaka and Okamoto 2021). The initial reduction in mental healthcare use in many countries are plausibly explained by limited access, as the healthcare sector was urged to reduce face-to-face patient contacts and patients being hesitant to seek care because of the infection risks. By comparing students in closed upper-secondary schools with students in open lower-secondary schools during the same time-periods, we effectively hold constant factors such as access to and the capacity of the healthcare system, the consequences of other NPIs, as well as other direct and indirect effects of the pandemic.

We study the probability of an adolescents being in contact with specialized psychiatric care or hospital or being prescribed a psychotropic drug in April-June 2020 compared to the same period in 2019. To investigate the consequences of school closures in the medium term we study how healthcare contacts develop in the fall and winter of 2020-2021 when school opened again. We investigate whether responses depend on sex, family socioeconomic status and whether the student was in contact with mental health care services earlier in the academic year. As the pandemic might induce a substitution between different points of care contacts, we also provide evidence on which types of care that are affected.

## Methods

### School closures and other disease containment measures in Sweden

On March 18, 2020 upper-secondary schools moved to online instruction while schools for younger students remained open until the summer break in mid-June. After the summer break upper-secondary schools reopened for in-person instruction but the possibility of partially using remote instruction remained. Most school relied fully on in-person teaching but some upper-secondary schools had students alternate between remote and in-person classes in order to reduce congestion at the schools and in public transport. In the beginning of the fall term, 80% of the schools taught at the premises and 20% had 10-40% of the classes online (Swedish Schools Inspectorate, 2020). Upper-secondary schools moved back to online instruction on December 7, 2020 to January 24 2021. Thereafter, upper-secondary schools were required to give each student at least 20% of the classes in person at schools (PHA, 2021). Students, who for different reasons were regarded to benefit from in person teaching, were generally allowed into the school premises (Swedish National Agency for Education, 2021b). As the infection rates decreased in the spring of 2021, schools gradually increased in-person schooling (Swedish National Agency for Education 2021 a,b,c).

Lower-secondary schools remained open throughout the spring and fall of 2020. From January 2021, lower-secondary schools were allowed to use remote teaching if local conditions so required. A survey by the Swedish Teachers’ Union (2021) in February 2021 reports that 88% of their lower-secondary members were to some extent teaching students in-person, while 49% were to some extent teaching without students in the classroom. Remote teaching decreased in the following months (Swedish National Agency for Education 2021 a,b,c).

We primarily study outcomes in April-June 2020 when upper-secondary schools were fully closed while lower-secondary schools remained open. We also provide results for July-December 2020, when upper secondary school were largely open, and January-March 2021, when both upper- and lower-secondary schools were partially open, although lower-secondary schools to a much larger extent.

In an international comparison, containment measures within Swedish schools were mild (Guthrie, 2020). Measures consisted of: enhanced information and facilities for promoting hand washing and disinfection; if possible, increased distance in classrooms and dining halls; avoidance of large gatherings and close contact between staff and students, and enhanced cleaning of heavily exposed areas. There were no recommendations or encouragement to use facemasks, no reduction in class size, and no targeted testing or quarantining of students. About the same time as upper-secondary schools closed, The Swedish Public Health Agency issued several other recommendations to reduce the spread of the virus such as: a ban against public gatherings above 50 persons; instructions to restaurants and bars to increase distance between guests; recommendations to stay at home if sick; work from home if possible and avoid unnecessary traveling. The containment measures where nation-wide and affected everyone in Sweden. In particular, lower- and upper-secondary students are likely to have been similarly affected by these measures as well by the pandemic in general. Since lower-secondary students attended school essentially as normal, comparing their use of mental healthcare services to that of upper-secondary students gives a credible estimate of the impact of school closures.

### Data and study population

The study population consists of all upper- and lower-secondary students in the academic years 2015/16 to 2020/21. The database used in this study is part of the research program “Covid-19 in Sweden: Infection tracing, control and effects on individuals and society” at Stockholm University. In the database, personal identifiers allow for linkages between different registries and families. Information on school year, sex and age as well as information on family and parental characteristics are originally from registers held by Statistics Sweden and information on health care use are from register data held by National Board of Health and Welfare. We identify all healthcare contacts at hospitals or doctors at specialist psychiatric care with diagnoses within chapter F in the ICD10 classification system - Mental, behavioral and neurodevelopmental disorder - and prescriptions of drugs for insomnia (ATC-code N05), attention deficit hyperactivity disorder (ADHD) (ATC-code N06B and C02AC02) and antidepressants (ATC-Code N06A). The main outcome variable is an indicator variable taking the value one if the student had any contact with a hospital or doctor at specialized psychiatric care for mental health issues, or was prescribed a drug for depression, insomnia or ADHD during each respective period. A drawback with the data is that it does not include information on contacts with primary healthcare. However, the prescription data include all information on all drugs prescribed in Sweden, also if prescribed by doctors working in primary care. We also study different diagnoses separately, if the contacts in specialist care were planned or not, and visits to emergency units and psychiatric emergency units with psychiatric diagnoses. As a test of whether access to healthcare decreased we study contacts with healthcare deemed unrelated to mental health and COVID-19. This measure includes all contacts with healthcare due to neoplasm (ICD10 C, D0-D4), diseases of skin and subcutaneous tissue (ICD10 L), endocrine, nutritional and metabolic diseases (ICD10 E) and diseases of the circulatory system (ICD10 I). See supplementary materials for data sources, detailed descriptions of variables and summary statistics of the sample (Table S1).

Ethical approval for the study was obtain from the Swedish Ethical Review Authority application 2020-06492.

### Statistical analysis

In the main analysis, we investigate the change in contact with mental health care services April-June 2020 compared to the same period in 2019. We also estimate the corresponding change compared to each year 2016-2018. Thus, we estimate the following difference-in-difference models using a linear regression (OLS):

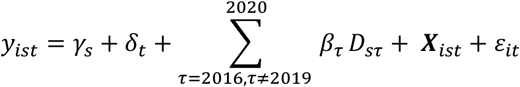

In the main analysis, the outcome *y*_*ist*_ is an indicator variable taking the value one if individual *i* was in contact with mental healthcare services or received a drug prescription for the causes described in the data section. *γ*_*s*_ is an indicator for whether the student attended upper-secondary school and *t* refers to the academic year. The estimates of β_*τ*_ are the differences between upper- and lower-secondary students for each academic year, compared to the reference year (2019) prior to the pandemic. **X** is a vector of individual- and parent characteristics. The variables included to adjust for potential compositional changes in the student population are: Indicators for student sex and birth month, whether the student immigrated to Sweden within the last 4 years or last 8 years, if born abroad or both parents are born abroad. We also include a range of parental characteristics reflecting the socio-economic position and family situation of the adolescents. Specifically, we include: 7 indicators of parental educational attainment; parental income percentile by year; age category and sex; indicators of unemployment and sickness absence the previous year; indicator of being a retiree; if the parents live together or are married; number of children under age 18 in the household; indicators for missing information on parents in registers.

The model is also estimated for the periods July-December 2020 and January-March 2021. We provide a sub-group analysis by splitting the sample by student sex, parental income (at least one parent in the 4^th^ income quartile), and if the student was in contact with mental healthcare services earlier in the academic year. To analyze if the change in use of mental healthcare services differ by school grade, we also estimate a model where we compare the outcome for students in each school grade 7-8, 10-12 relative to school grade 9 before and after schools closed (for specification see supplementary materials).

## Results

The use of mental health care services among adolescents has increased for several years in Sweden (The National Board of Health and Welfare, 2019). These trends are illustrated in Figure 1 for the April-June-period among upper- and lower secondary students. The figure displays the estimates from a linear regression of contacts with mental health services on year fixed effects for upper-respectively lower secondary students. Before the pandemic, use of mental health care services increased in tandem for lower- and upper-secondary students with about 5 cases per 1000 each year. In 2020, when upper-secondary schools were closed, contacts with mental healthcare services among lower-secondary students increased at the same rate as earlier years, whereas contacts among upper-secondary students remained at the 2019 level. These results suggest there is no general drop in contacts with mental health care services as lower-secondary students do not seems to be affected.

**Figure 1.**
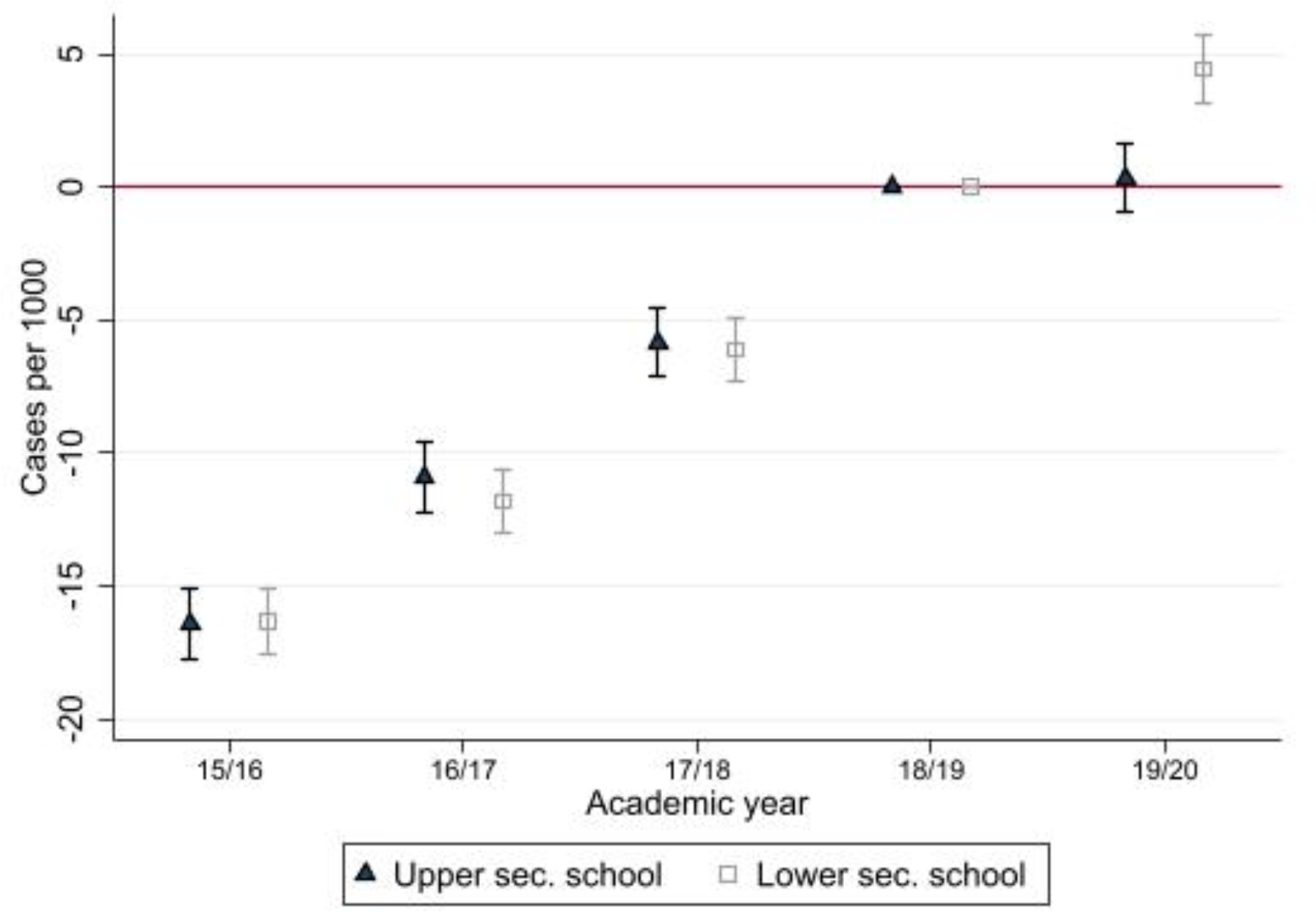
Trends in use of mental healthcare services. Note: Estimates from separate linear regressions for upper-secondary respectively lower-secondary students. 95% CI indicated.

To formally test whether upper-secondary students reduced use of mental health care services we estimate the difference-in-difference model specified above. Relative to the expected rate, upper-secondary students reduced their contacts with 3.71 [95% CI (CI95) -5.52 to -1.91] cases per 1000 in the period April-June 2020 (supplement materials Table S2). This implies a 4.36% decrease relative to the mean for upper-secondary students in 2019. Figure 2 shows the difference-in-difference estimates for each year rescaled to changes in percent. Contacts with mental healthcare services followed a similar trend for the two groups in the years preceding the pandemic. Thus, the change in use between upper- and lower-secondary students in 2020 is unlikely due to differences in long-term trends. Furthermore, the change in 2020 is not due to compositional changes in the student population, as the estimates are not affected by adjusting for background characteristics (supplementary materials Table S2).

**Figure 2.**
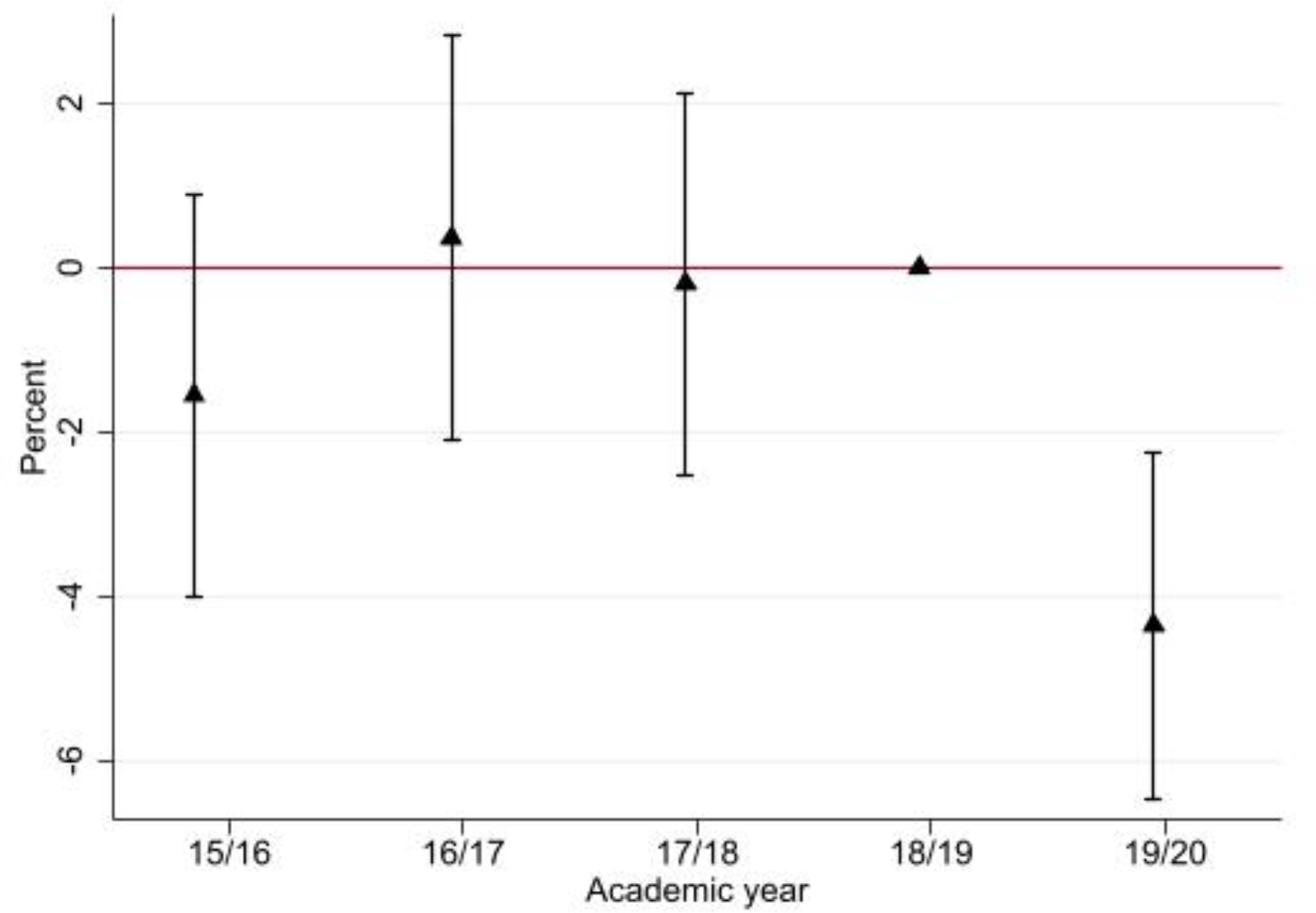
Mental healthcare use among upper-secondary students. Note: Difference-in-difference estimates from linear regression rescaled to percent. Lower-secondary students as control group for upper-secondary students. Academic year 2018/19 is the reference period. 95% CI indicated.

Next, we investigate if use of mental health care services in April-June 2020 changed differently depending on the students’ school grade. Figure 3 shows estimated changes, rescaled to percent, of contacts with mental healthcare services April-June 2020 for each school grade relative to grade 9 (see supplementary materials Table S3). For grades 7 and 8, the estimates are small and statistically insignificant. This is expected as all lower-secondary students had in-person schooling during the period. For grade 10, the first year of upper-secondary school, the estimate is negative but not statistically significant (-1.93; CI95 [-4.41 to 0.55]). The negative estimates are larger and statistically significant for grade 11 (-4.32; CI95 [-6.86 to -1.79]) and 12 (-3.70; CI95 [-6.24 to 1.16]). Compared to the mean, the decrease is 2.11% for grade 10, 5.06% for grade 11, and 4.86% for grade 12.

**Figure 3.**
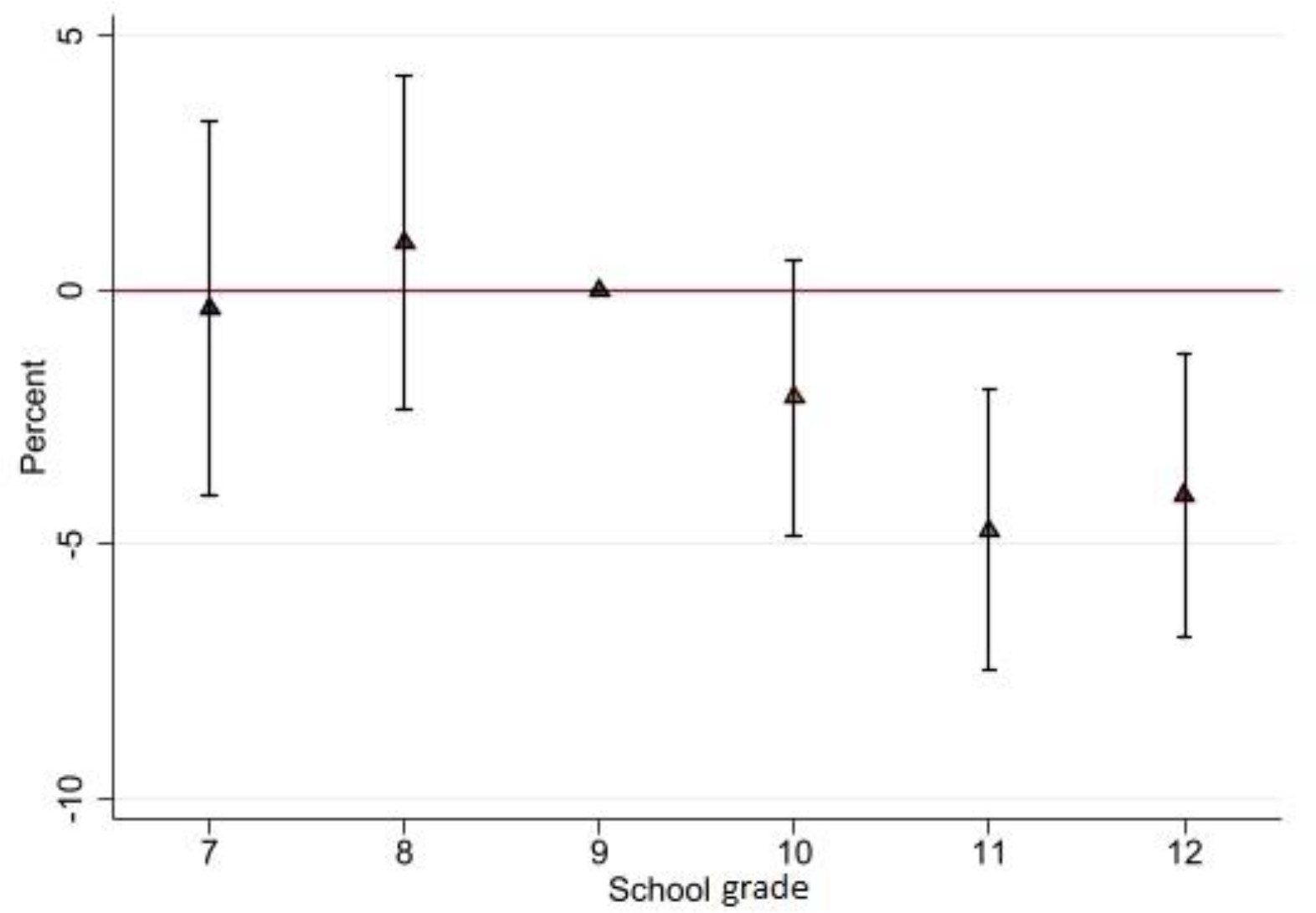
Mental healthcare use among students in school grades 7-12. Note: Difference-in-difference estimates from linear regression rescaled to percent. Academic year 2019/20 relative to mean prior to the pandemic. School grade 9 is the reference category. 95% CI indicated.

Figure 4 displays the difference-in-difference estimates scaled to changes in percent for 2020 compared to the reference year 2019 for different sub-populations (see complete results in supplementary materials Table S2). The negative estimate is larger for men/boys, -6.30% (-4.26; CI95 [-6.65 to -1.88]) than for girls/women -3.07% (-3.22; CI95 [-5.95 -0.50]). The change is larger among students from higher income families -6.81% (-5.93; CI95 [-8.73 to -3.13]) than lower income families -1.89% (-1.64; CI95 [-4.07 to 0.78]). Dividing the population on mental healthcare use earlier in the academic year shows that students who had received no such care account for the entire reduction -16.70% (-2.40; CI95 [-3.19 to -1.61]).

**Figure 4.**
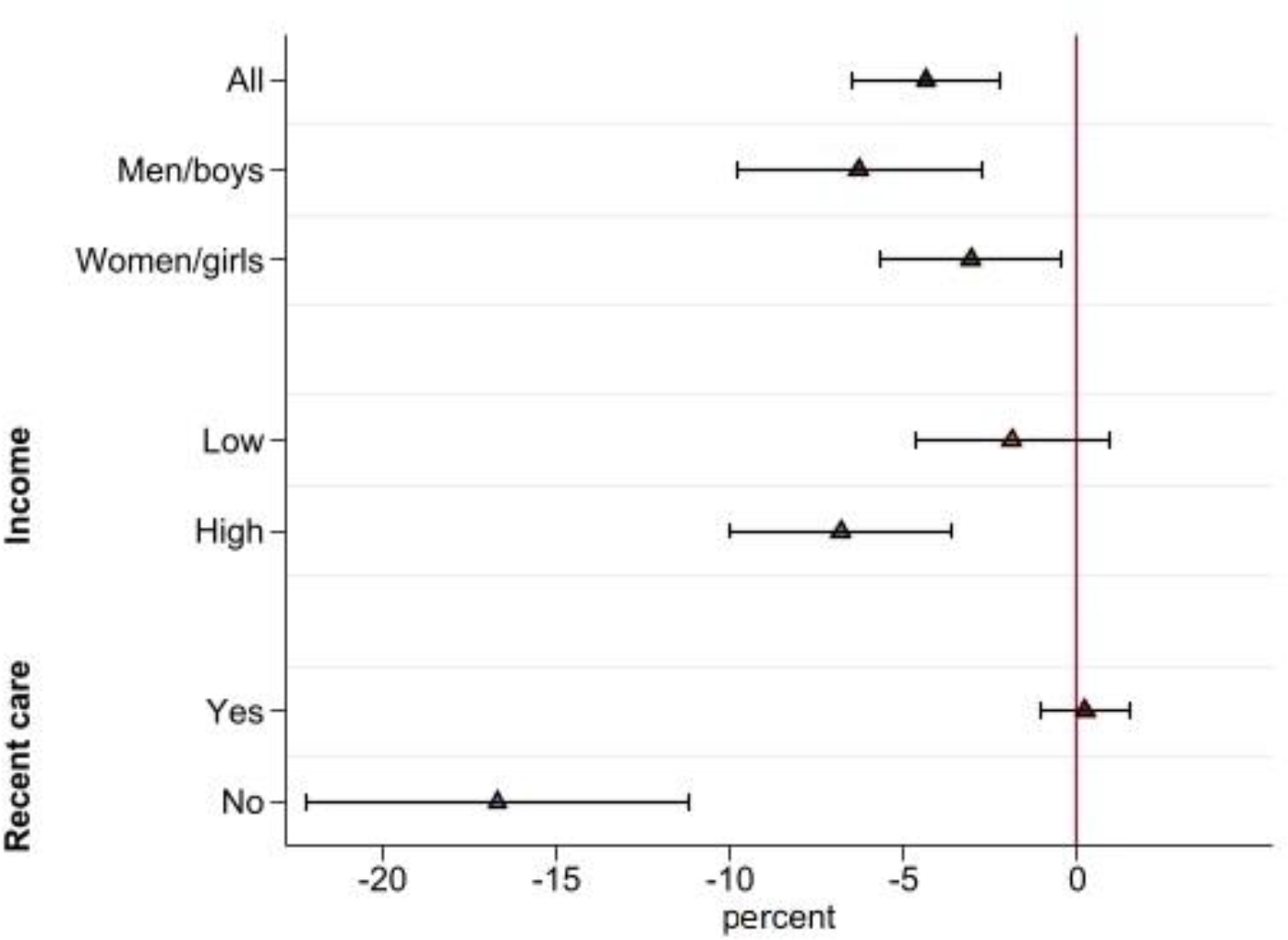
Estimates for sub-groups. Note: Difference-in-difference estimates by sex, parental income, and if the person had contact with health care earlier during the academic year. Estimates from separate linear regressions rescaled to percent. 95% CI indicated.

Results for different sub-diagnoses and psychotropic drugs are presented in **Fel! Hittar inte referenskälla**. in the supplementary materials. Estimates show reductions mainly for depression and anxiety. Healthcare contacts with diagnoses for depression and anxiety is reduced with -7.37% (-1.49; CI95 [-2.36 to -0.63]) and prescriptions of antidepressants with - 4.70% (-1.80; CI95 [-2.93 to -0.68]). To analyze if there is a general reduction in accessibility of healthcare, specific for upper-secondary students, we study the change in diagnoses likely unrelated to mental health and COVID-19. There is no indication this is the fact as the estimate is small in magnitude and statistically insignificant (0.06; CI95 [-0.99 to 1.10]).

After the summer break 2020, upper-secondary schools opened for in-person instruction. With local exceptions, most upper-secondary schools remained open until December 7 when they closed because of high infection rates. After the winter break, upper-secondary schools had some level of in-person instruction between January-March 2021. In these periods, contacts with mental healthcare services increased both among upper- and lower-secondary students (supplement material Figure S1). Using lower-secondary students as the control group, we estimate the difference-in-difference model for the periods July-December and January-March. The results displayed in Figure 5 (coefficients in supplementary material Table S5) show persistent reductions in use of mental healthcare services among upper-secondary students in the fall of 2020 with -3.55% (-3.75; CI95 [-5.69 to -1.81] and winter/spring of 2021 with - 5.23% (-4.79; CI95 [-6.64 to -2.95]).

**Figure 5.**
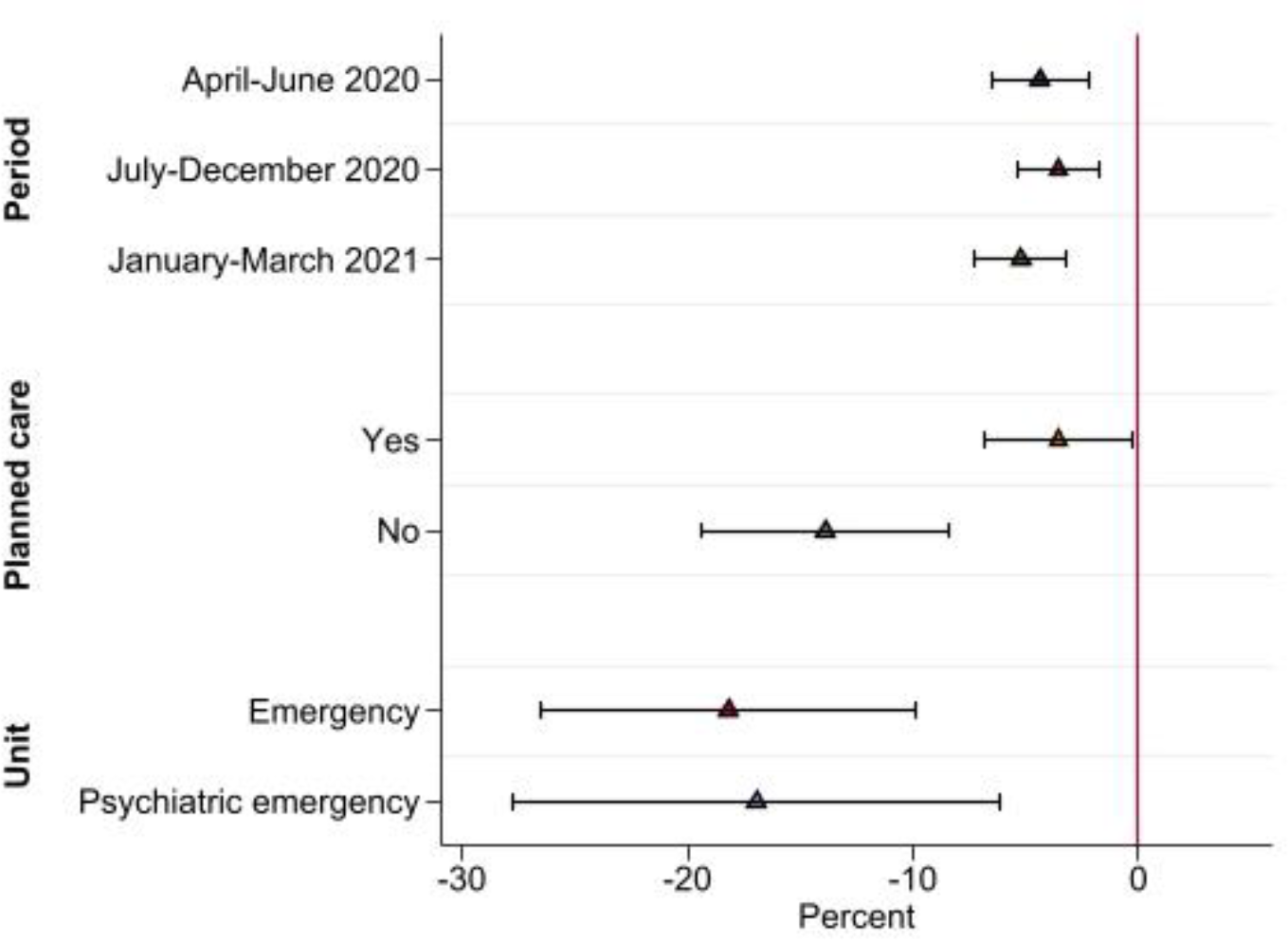
Estimates by time-periods and types of care. Note: Difference-in-difference estimates from separate linear regressions rescaled to percent. 95% CI indicated.

Prescriptions of antidepressants display a small decrease in the spring of 2020 among upper-secondary students. In the fall of 2020 and winter/spring of 2021, prescriptions increased but at a lower rate than among lower-secondary students (supplementary material Figure S2). Healthcare contacts with diagnoses for depression and anxiety decreased among upper-secondary students in April-June 2020, remained constant July-December 2020, and July-Mars 2021, compared to the same periods the previous year. For this outcome, there is also a visible deviation from trend among lower-secondary students during April-June 2020, indicting a general reduction in contacts for depression and anxiety. However, throughout all periods, the reduction is larger among upper-secondary students (supplement material Figure S3).

The closure of upper-secondary schools between March-June 2020 potentially reduced access to psychiatric care through schools’ healthcare facilities, which could imply the reduction in use is due to reduced asses to healthcare. If access through school facilities is reduced, this would plausibly shift upper-secondary students (relative to lower-secondary students) towards unplanned and emergency visits to psychiatric care facilities. However, compared to lower-secondary students the relative decrease in contacts is substantially larger for unplanned - 13.88% (-1.65; CI95 [-2.31 to -1.00]) than for planned -3.56% (-1.39; CI95 [-2.66 to-0.11]) contacts during April-June 2020. In particular, there is a large decrease in contacts with emergency or psychiatric emergency units among upper-secondary students compared to students in lower-secondary schools -16.70% (-0.38; CI95 [-0.62 to -0.14]). Also during the fall 2020 and winter/spring 2021, the relative decrease is substantially larger for unplanned contacts and contacts with emergency care units (Supplementary Materials Table S5 and Table S6). These results are inconsistent with the patterns of substitution between providers that we would expect if reduced access through school facilities lies behind the main results.

## Discussion

This study analyses the impact of school closures on use of mental healthcare services among upper-secondary students. In the wake of the pandemic, Sweden closed upper-secondary schools from March 18 until the summer break in mid-June while keeping schools at lower levels open. We exploit this partial school closure to identify the impact of school closures on mental healthcare use by comparing upper-(grades 10-12) and lower-(grades 7-9) secondary students, before and after the school closure. The results show that upper-secondary students reduced their use of mental healthcare services by 4.36 percent compared to lower-secondary students. This lower utilization persists up until the end of the study period in March 2021. The analysis shows reductions in all groups, although somewhat larger among boys/men, students from higher-income families, and in school grades 11 and 12. The largest reductions are seen in contacts for depression and anxiety, and prescriptions for antidepressants.

The analysis shows that the reduction was due to reduced use among students with no contact with healthcare for mental health issues earlier in the academic year. This group was 17 percent less likely to be in contact with mental healthcare services during the initial closure. There is little evidence suggesting this may be due to limited access to care. In contrast, three results speak against such an interpretation. First, the analysis shows the largest relative reductions in unplanned contacts with psychiatric care facilities and in contacts with psychiatric emergency units. If upper-secondary students did not get the care they needed via normal channels, we would instead expect some substitution towards emergency units. Second, contacts with mental healthcare services did not increase when upper-secondary schools opened in the fall of 2020 or during the winter/spring of 2021 when upper-secondary schools were partially open. This suggests that there was no excess demand due to previously unmet needs. Third, healthcare contacts for issues unrelated to mental health and COVID-19 did not change differently between upper- and lower-secondary students. If reduced access for upper-secondary students is to explain the results presented, it would then have to be quite specific to mental healthcare. Further, while many countries have reported extensive disruptions in mental health services (WHO 2021), specialized psychiatric care in Sweden seems to have managed well by moving consultations online (National Board of Health and Welfare 2021).

The main contribution of this study is that we compare the outcomes of students facing in-person and remote schooling during the same time-periods, thereby adjusting for factors that affect both groups similarly. Thus, we do not estimate the total effect of the pandemic but isolate the impact of moving to remote instruction. In line with studies from other countries (Carr et al. 2021; Evensen et al. 2021; Ougrin 2020), we find healthcare contacts with diagnoses for depression and anxiety and prescriptions of antidepressants to decline relative to the increasing trend in April-June 2020 also among lower-secondary students. This decline is, however, small and for the periods July-December 2020 and January-March 2021, mental healthcare use among lower-secondary students increase at the same rate as in the previous years. Again, this suggest access to mental health services was not severely affected during the pandemic.

Should the results be interpreted as if remote learning improves mental health? As discussed, psychiatric care has been accessible during the pandemic and there are no signs of large unmet needs among upper-secondary students. It is important to take the Swedish context into account when interpreting the results. In an international comparison, the restrictions in the Swedish society have been mild. Although there were recommendations against travelling, meeting with elderly, and socializing with more than 8 persons there were, for example, no stay-at-home orders and restaurants and gyms largely remained open. Thus, it was possible for upper-secondary students to see friends and engage in social activities outside of school. In addition, the full school closure lasted less than 3 months. The potential negative consequences of remote learning are likely larger when students are isolated for longer time periods. Another feature with the Swedish setting is that internet penetration is high and the conditions for remote learning are rated among the best in the world (OECD 2021b). Reports also suggest that the move to online teaching in upper-secondary school have worked relatively well, albeit with some problems (Swedish National Agency for Education 2020). As in many countries, standardized tests were largely cancelled which may reduce stress but as tests in lower-secondary school were also cancelled, this alone cannot account for the differences between lower- and upper-secondary students. Among US high school students, there is evidence that the disruption of in-person schooling led to a substantial reduction of bullying, including cyberbullying (Bacher-Hicks et al, 2021). While we are not aware of similar evidence from Sweden, it is another potential channel through which remote learning can have a positive impact on mental health.

Ideally, this study would include self-reported surveys of mental health to understand if the reduction can be interpreted improvement in mental health. No such information is available, however. The one study of adolescents in Sweden studying mental health during the pandemic using survey data finds no change in mental health among 16-year-olds (Chen et al 2021). This is in line with our finding of no change in mental health among lower-secondary students (aged 14-16). Survey evidence on adolescents’ emotional and behavioral problems from other countries during the pandemic, that attempts to adjust for selection as well as time and age trends, find both deteriorating (e.g Thorisdottier et al. (2021); Luijten et al. (2021); Rogers et al. (2021) and unaffected mental health (e.g Munasinghe et al. (2020); Hafstad et al. 2021).

This study has some limitations. The outcome measure includes contacts with hospitals and specialized psychiatric care, and prescriptions from any prescriber of drugs in Sweden. However, it does not include information on contacts with primary care or personnel working with healthcare in schools. School closures may have reduced access to school nurses and counselors who may refer students to specialized mental healthcare services. Thus, although the evidence presented suggests otherwise, we cannot entirely rule out this explanation. Another limitation is that other disease containment measures, such as restrictions on nightlife and travels, may have differently affected on upper- and lower secondary students. A natural limitation is the focus on immediate and medium-term effects. School closures may have reduced learning and opportunities for social interactions that can have detrimental long-term implications.

## Data Availability

Data is available from Statistics Sweden and the National Board of Welfare following an ethical review.

## Supplementary Materials

### Methods and Material

#### Data

The estimation sample is constructed using the Student Register held by Statistics Sweden. The sample consists of all students in upper- and lower secondary school the academic years 2015/16-2020/21. Students are connected to their parents and to information on healthcare use through a personal identifier. Specifically, students are connected to their biological or adoptive parents using the Multi-Generational register. Information on demographic and socio-economic variables of the parents are taken from the Longitudinal integrated database for health insurance and labor market studies (LISA) maintained by Statistics Sweden. Data on contacts with doctors in specialized psychiatric care and hospital visits is taken from the Patient registers, and drug prescriptions from the Drug register. Both these registers are held by the National Board of Health and Welfare.

Definitions of outcome variables are described in the main text. Most control variables are self-explanatory but the income measure renders a description. The measure of parental income is based on individual disposable income and constructed as follows: We use the average income for the years 2015-2019 and percentile rank each individual by birth cohort and sex. Note that the rank measure is constructed using all individuals in Sweden. For newly immigrated individuals we only use the income after immigration. When dividing the sample by income, a student is coded as “high income” if any of the parents has an income rank in the fourth quartile.

Summary statistics of the main outcome variable, sex, foreign background and parental educational attainment and income percentile are presented in Table S1. The left panel shows values for the academic years 2015/16 to 2020/21 for lower-secondary students and the right panel for upper-secondary students. As can be seen in the table, the use of mental healthcare services increases over time for both groups prior to the pandemic. When upper-secondary schools closed, mental healthcare use among upper-secondary students in April-June 2020 decreased from 85.21 per 1000 to 84.83 per 1000 whereas use among lower-secondary students increased from 75.00 to 78.45 students per 1000. Inspecting the background variables show no drastic change over time. In particular, judging by these variables there is no change in the composition of students in the academic years 2019/20 and 2020/21 that could explain changes in use of mental health services. Students in upper-secondary school are slightly more likely to be male, have foreign background and have parents without university education and lower income. The reason is that students without complete school leaving certificate from lower-secondary school begin preparatory programs in upper-secondary school to attain the requirements in core subjects. These students then stay more than one year in grade 10.

#### Statistical method

The estimates for the year effects displayed in Figure 1 are retrieved from estimating the model using regression (OLS) separately for lower- and upper secondary school students:

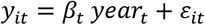

Where *y*_*it*_ is the outcome for student *i* in year *t*.

The difference-in-differences estimates displayed in Figure 3 and Table S3 are retrieved by estimating the following model using linear regression (OLS):

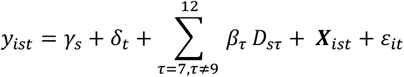

Where *y*_*it*_ is the main outcome variable, *t* indicates before or after schools closed and *s* is student school grade. The estimate *β*_*τ*_ show the difference between school grades s= 7-8 and 10-12, compared to the reference school grade 9, in April-June 2020 compared to the average April-June 2016-2020.

## Figures

**Figure S1.**
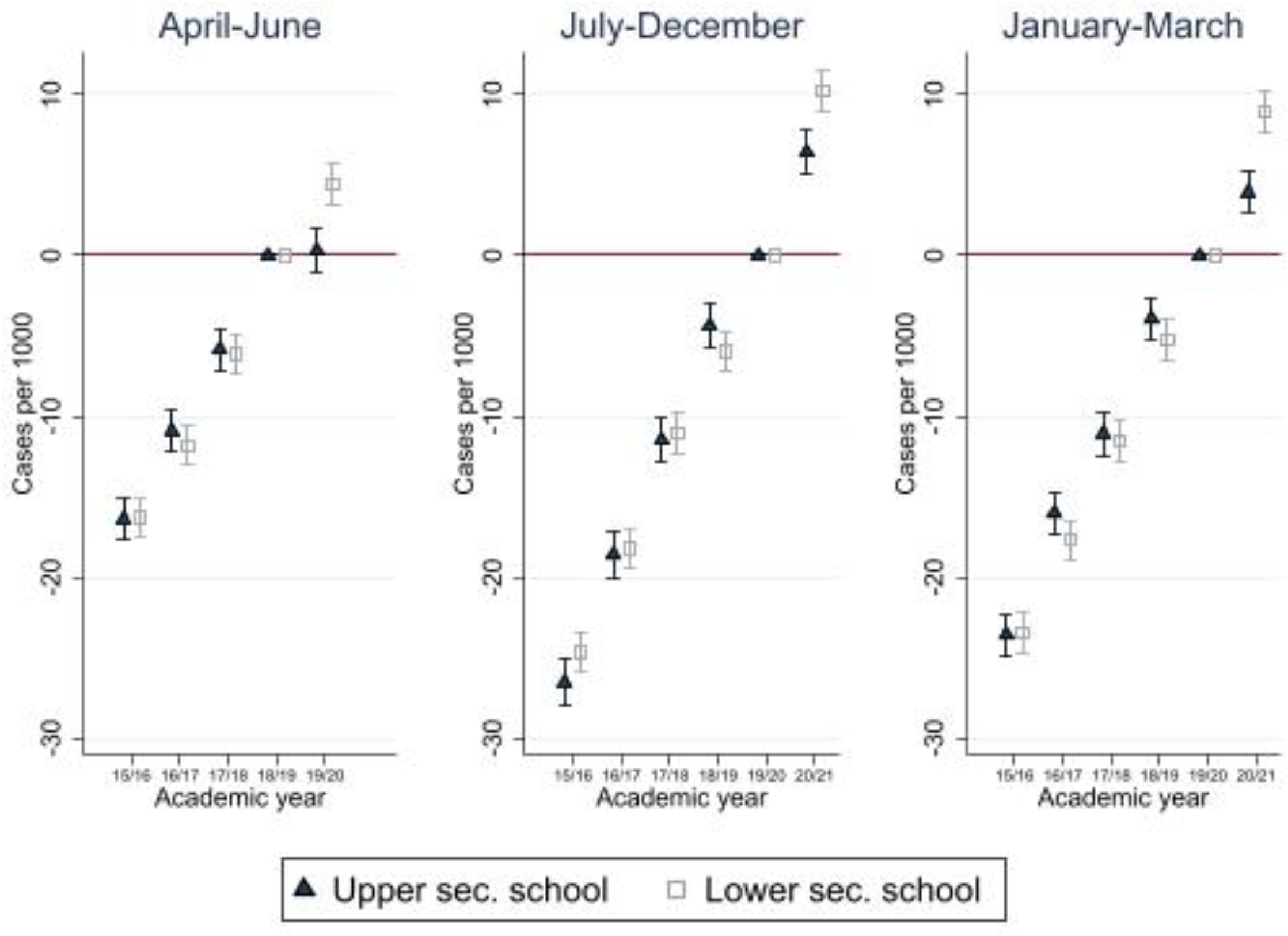
Trends in use of mental healthcare services. Note: Estimates from separate linear regressions for upper-secondary and lower-secondary students. 95% CI indicated.

**Figure S2.**
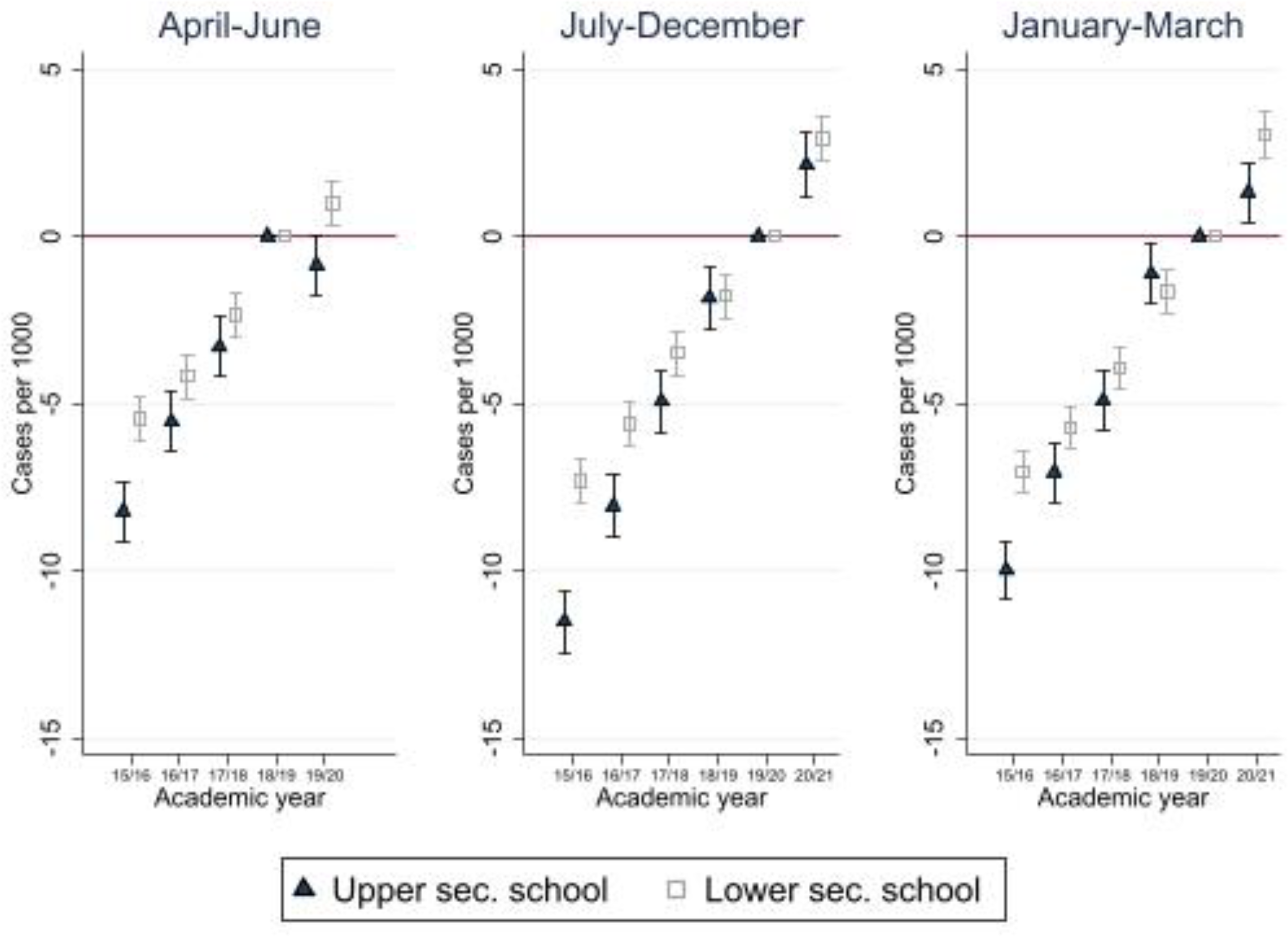
Prescriptions of antidepressants. Note: Estimates from separate linear regressions for upper-secondary and lower-secondary students. 95% CI indicated.

**Figure S3.**
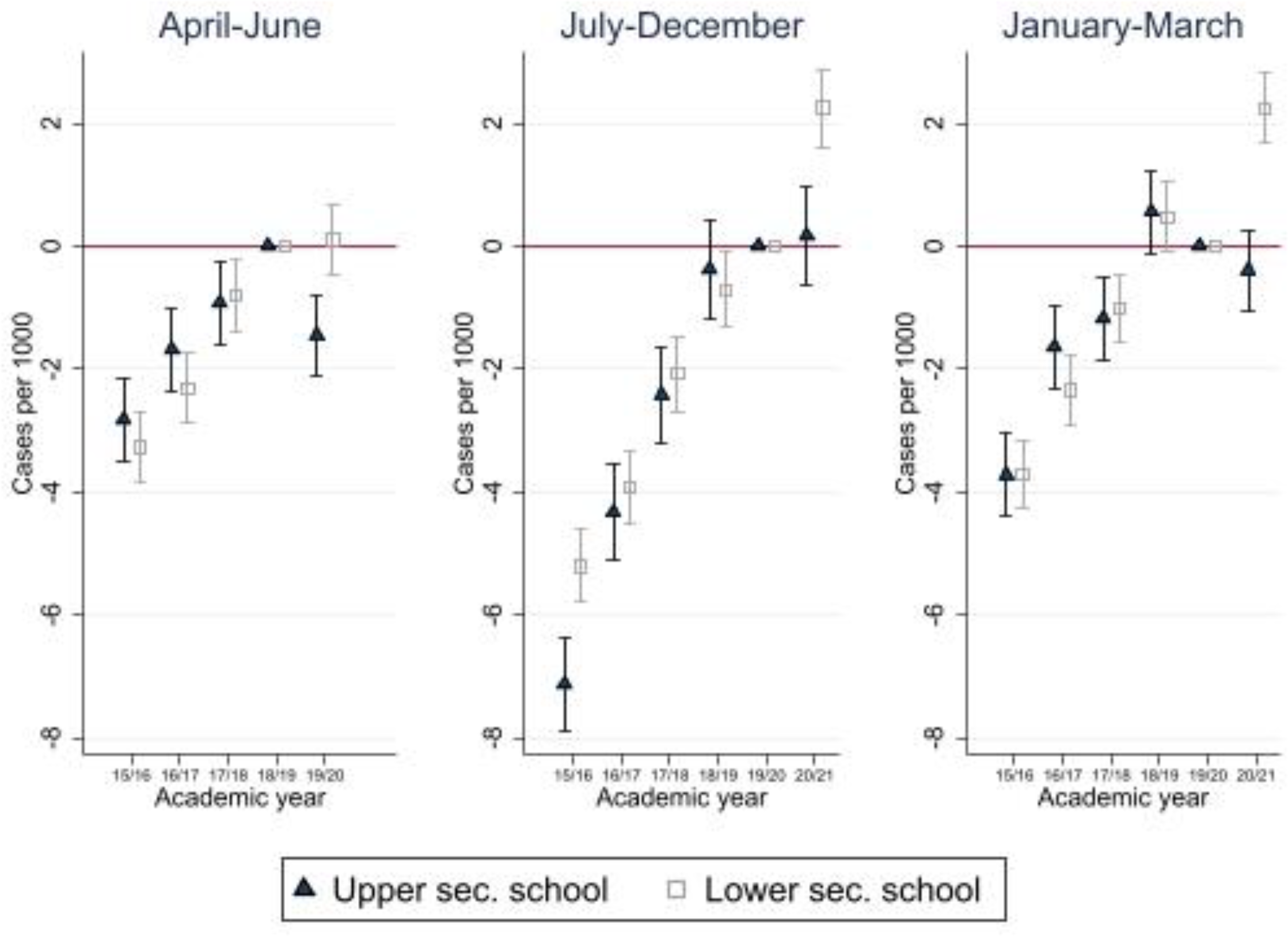
Healthcare contacts for depression or anxiety. Note: Estimates from separate linear regressions for upper-secondary and lower-secondary students. 95% CI indicated.

## Tables

**Table S1.**
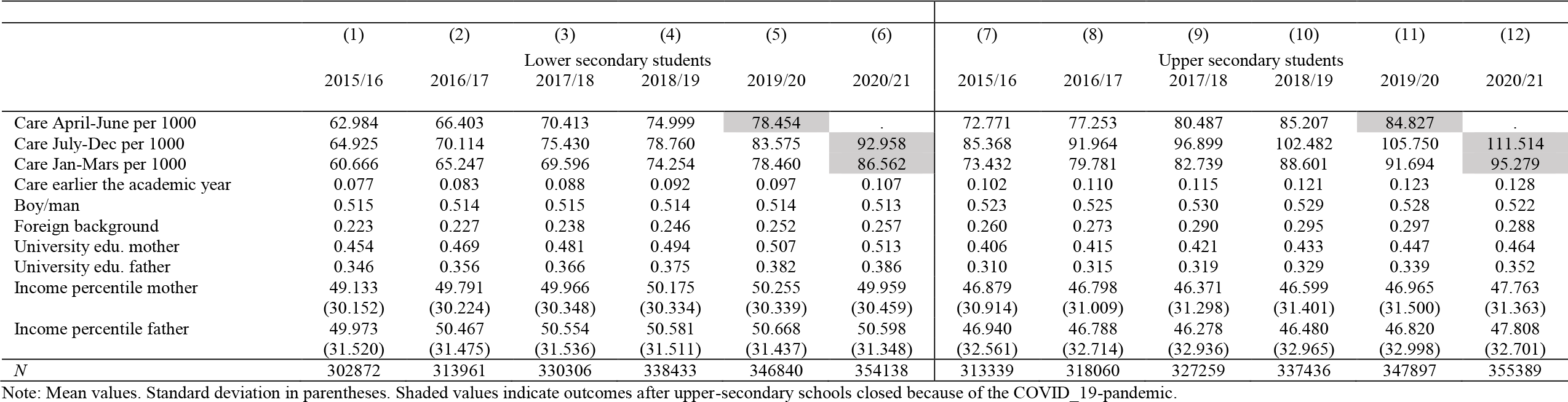
Summary statistics

**Table S2.** Difference-in-difference estimates of psychiatric care or prescriptions. Main results and results for subgroups

|  | (1)<br>No controls | (2)<br>Controls | (3)<br>Men | (4)<br>Women | (5)<br>Low income | (6)<br>High income | (7)<br>Recent contact | (8)<br>No recent contact |
| --- | --- | --- | --- | --- | --- | --- | --- | --- |
| | $\beta$<br>[95% CI]<br>(p-value) | $\beta$<br>[95% CI]<br>(p-value) | $\beta$<br>[95% CI]<br>(p-value) | $\beta$<br>[95% CI]<br>(p-value) | $\beta$<br>[95% CI]<br>(p-value) | $\beta$<br>[95% CI]<br>(p-value) | $\beta$<br>[95% CI]<br>(p-value) | $\beta$<br>[95% CI]<br>(p-value) |
| Upper sec × 2016 | -0.422<br>[-2.225,1.381]<br>(0.647) | -1.133<br>[-2.915,0.649]<br>(0.213) | 1.826<br>[-0.560,4.213]<br>(0.134) | -4.486***<br>[-7.154,-1.818]<br>(0.001) | 0.398<br>[-2.001,2.797]<br>(0.745) | -2.859*<br>[-5.616,-0.103]<br>(0.042) | -7.977<br>[-18.495,2.541]<br>(0.137) | 0.468<br>[-0.352,1.287]<br>(0.263) |
| Upper sec × 2017 | 0.640<br>[-1.175,2.455]<br>(0.489) | 0.256<br>[-1.537,2.049]<br>(0.780) | 0.024<br>[-2.359,2.407]<br>(0.984) | 0.489<br>[-2.216,3.193]<br>(0.723) | 2.739*<br>[0.321,5.157]<br>(0.026) | -2.776*<br>[-5.550,-0.001]<br>(0.050) | 4.660<br>[-5.600,14.921]<br>(0.373) | 0.123<br>[-0.695,0.941]<br>(0.768) |
| Upper sec × 2018 | -0.136<br>[-1.954,1.682]<br>(0.883) | -0.151<br>[-1.946,1.643]<br>(0.869) | -1.068<br>[-3.451,1.316]<br>(0.380) | 0.907<br>[-1.804,3.617]<br>(0.512) | 1.278<br>[-1.141,3.697]<br>(0.301) | -1.661<br>[-4.456,1.135]<br>(0.244) | 2.492<br>[-7.470,12.454]<br>(0.624) | 0.608<br>[-0.198,1.415]<br>(0.139) |
| Upper sec × 2020 | -3.836***<br>[-5.661,-2.010]<br>(0.000) | -3.714***<br>[-5.515,-1.913]<br>(0.000) | -4.262***<br>[-6.649,-1.876]<br>(0.000) | -3.223*<br>[-5.948,-0.498]<br>(0.020) | -1.642<br>[-4.068,0.784]<br>(0.185) | -5.933***<br>[-8.733,-3.133]<br>(0.000) | 1.625<br>[-7.965,11.216]<br>(0.740) | -2.402***<br>[-3.193,-1.611]<br>(0.000) |
| Mean dep var upper sec 2019 | 85.21 | 85.21 | 67.69 | 104.90 | 86.78 | 87.07 | 600.34 | 14.38 |
| Effect in % | -4.50 | -4.36 | -6.30 | -3.07 | -1.89 | -6.81 | 0.27 | -16.70 |
| R2 | 0.001 | 0.025 | 0.025 | 0.028 | 0.035 | 0.015 | 0.034 | 0.003 |
| N | 3,276,398 | 3,276,398 | 1,706,036 | 1,570,362 | 1,867,375 | 1,338,745 | 331,388 | 2,945,010 |

Note: Results from linear regression model specified in main text. Reference year 2019. Specifications presented in (2)-(8) adjust for: sex, birth month, newly immigrated to Sweden, parental foreign background, income, education, income from social security systems, family size and if the biological parents live in the same household.

**Table S3.**
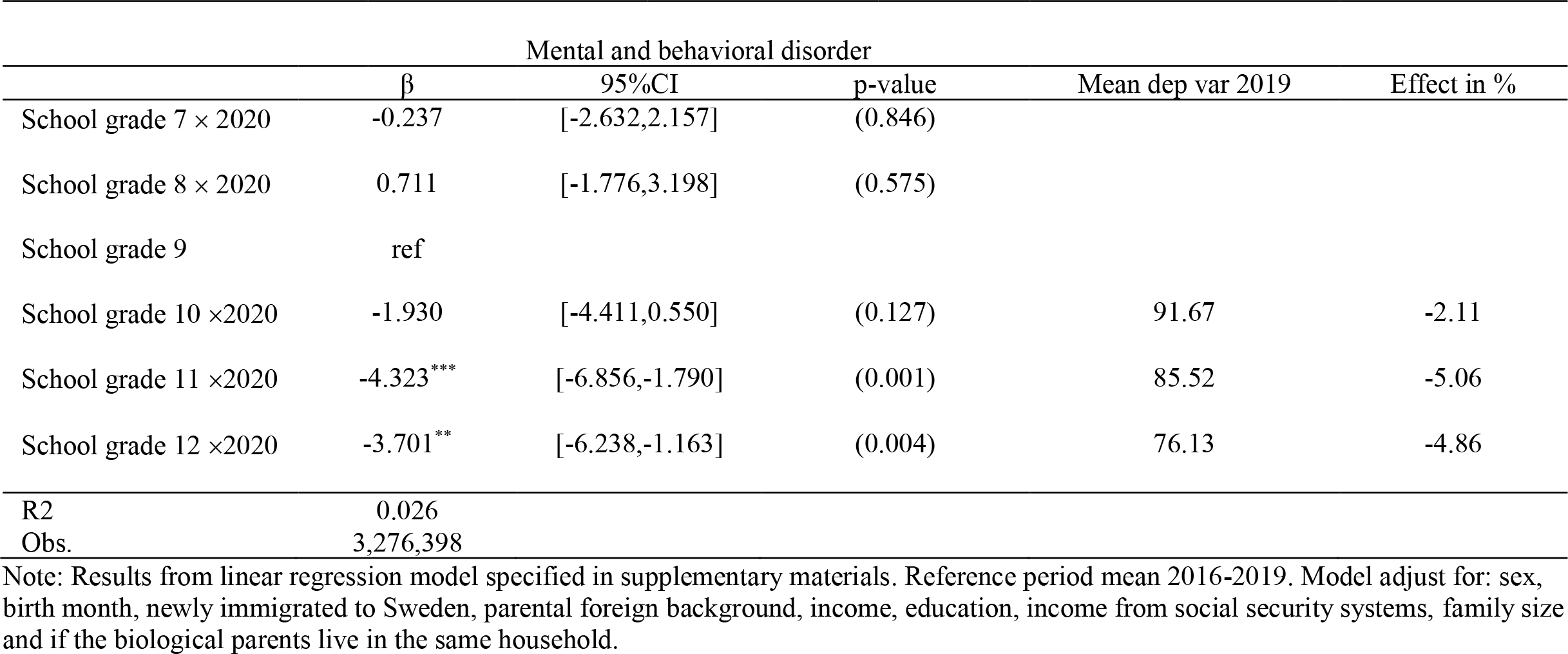
Difference-in-difference estimates by school grade

|  | Mental and behavioral disorder |  |  | Mean dep var 2019 | Effect in % |
| --- | --- | --- | --- | --- | --- |
| | $\beta$ | 95%CI | p-value | | |
| School grade 7 × 2020 | -0.237 | [-2.632,2.157] | (0.846) | | |
| School grade 8 × 2020 | 0.711 | [-1.776,3.198] | (0.575) | | |
| School grade 9 | ref |  |  |  |  |
| School grade 10 × 2020 | -1.930 | [-4.411,0.550] | (0.127) | 91.67 | -2.11 |
| School grade 11 × 2020 | -4.323*** | [-6.856,-1.790] | (0.001) | 85.52 | -5.06 |
| School grade 12 × 2020 | -3.701** | [-6.238,-1.163] | (0.004) | 76.13 | -4.86 |
| R2 | 0.026 |  |  |  |  |
| Obs. | 3,276,398 |  |  |  |  |

Note: Results from linear regression model specified in supplementary materials. Reference period mean 2016-2019. Model adjust for: sex, birth month, newly immigrated to Sweden, parental foreign background, income, education, income from social security systems, family size and if the biological parents live in the same household.

**Table S4.**
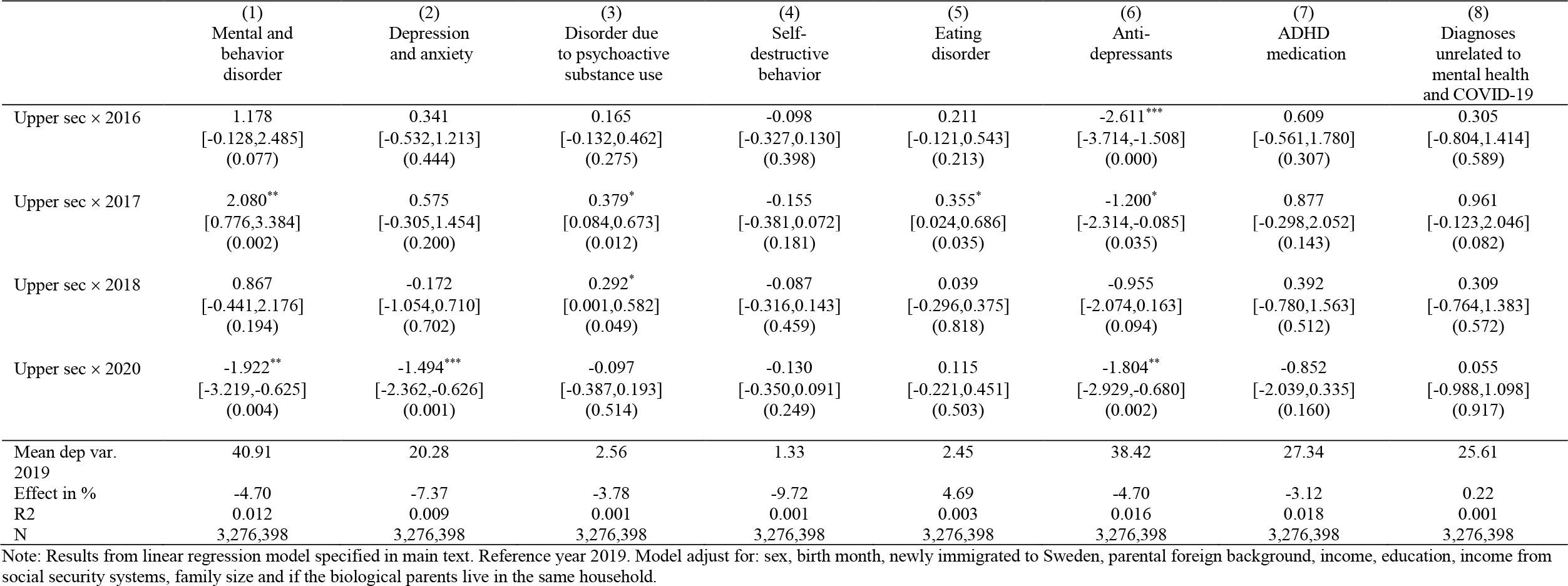
Difference-in-difference estimates. Subdiagnoses and perscriptions of antidepressants and ADHD drugs

**Table S5.** Difference-in-difference estimates for outcomes July-March, planned and unplanned specialist psychiatric care

|  | (1)<br>Jul-Dec 2020 | (2)<br>Jan-Mar 2021 | (3)<br>Planned care<br>Apr-Jun | (4)<br>Planned care<br>Jul-Dec | (5)<br>Planned care<br>Jan-Mar | (6)<br>Unplanned care<br>Apr-Jun | (7)<br>Unplanned care<br>Jul-Dec | (8)<br>Unplanned care<br>Jan-Mar |
| --- | --- | --- | --- | --- | --- | --- | --- | --- |
| Upper sec × 2016 | -2.509**<br>[-4.387,-0.632]<br>(0.009) | -1.300<br>[-3.088,0.489]<br>(0.154) | 1.380*<br>[0.087,2.672]<br>(0.036) | -1.307<br>[-2.811,0.197]<br>(0.089) | 0.938<br>[-0.342,2.218]<br>(0.151) | 0.249<br>[-0.451,0.949]<br>(0.485) | 1.021*<br>[0.117,1.925]<br>(0.027) | 1.313***<br>[0.637,1.989]<br>(0.000) |
| Upper sec × 2017 | -0.815<br>[-2.711,1.081]<br>(0.399) | 0.788<br>[-1.020,2.595]<br>(0.393) | 1.936**<br>[0.650,3.223]<br>(0.003) | 0.295<br>[-1.217,1.807]<br>(0.702) | 2.020**<br>[0.719,3.320]<br>(0.002) | 0.560<br>[-0.134,1.255]<br>(0.114) | 0.564<br>[-0.327,1.456]<br>(0.215) | 0.905**<br>[0.232,1.578]<br>(0.008) |
| Upper sec × 2018 | -0.870<br>[-2.773,1.032]<br>(0.370) | -0.230<br>[-2.040,1.579]<br>(0.803) | 0.692<br>[-0.597,1.981]<br>(0.293) | -0.417<br>[-1.933,1.099]<br>(0.590) | 0.066<br>[-1.224,1.356]<br>(0.920) | 0.990**<br>[0.301,1.679]<br>(0.005) | 0.762<br>[-0.121,1.644]<br>(0.091) | 0.887**<br>[0.225,1.549]<br>(0.009) |
| Upper sec × 2019 | 1.391<br>[-0.519,3.301]<br>(0.153) | 0.986<br>[-0.837,2.810]<br>(0.289) | 0.000<br>[0.000,0.000]<br>(.) | 0.529<br>[-0.990,2.048]<br>(0.495) | 0.477<br>[-0.824,1.778]<br>(0.472) | 0.000<br>[0.000,0.000]<br>(.) | 0.907*<br>[0.031,1.784]<br>(0.042) | 0.262<br>[-0.397,0.922]<br>(0.435) |
| Upper sec × 2020 | ref | ref | -1.385*<br>[-2.663,-0.107]<br>(0.034) | ref | ref | -1.654***<br>[-2.305,-1.003]<br>(0.000) | ref | ref |
| Upper sec × 2021 | -3.749***<br>[-5.687,-1.811]<br>(0.000) | -4.793***<br>[-6.640,-2.946]<br>(0.000) |  | -2.902***<br>[-4.431,-1.374]<br>(0.000) | -3.034***<br>[-4.326,-1.743]<br>(0.000) |  | -2.011***<br>[-2.869,-1.153]<br>(0.000) | -1.668***<br>[-2.304,-1.032]<br>(0.000) |
| Mean dep var upper sec<br>2019 | 105.75 | 91.69 | 38.95 | 60.78 | 41.25 | 11.91 | 20.43 | 11.18 |
| Effect in % | -3.55 | -5.23 | -3.56 | -4.78 | -7.36 | -13.88 | -9.84 | -14.92 |
| R2 | 0.031 | 0.028 | 0.011 | 0.017 | 0.012 | 0.002 | 0.003 | 0.002 |
| N | 3,985,925 | 3,985,925 | 3,276,398 | 3,985,925 | 3,985,925 | 3,276,398 | 3,985,925 | 3,985,925 |

Note: Results from linear regression model specified in main text. Models adjust for: sex, birth month, newly immigrated to Sweden, parental foreign background, income, education, income from social security systems, family size and if the biological parents live in the same household. Reference period in (1), (4) and (7) are July-December 2019, in (2), (5) and (8) January-Mars 2021 and in (3) and (6) April-June 2019.

**Table S6.** Difference-in-difference estimates for outcomes emergency unit visits

|  | (1)<br>Emergency unit<br>Apr-Jun | (2)<br>Emergency unit<br>Jul-Dec | (3)<br>Emergency unit<br>Jan-Mar | (4)<br>Psychiatric emergency<br>unit Apr-Jun | (5)<br>Psychiatric emergency<br>unit Jul-Dec | (6)<br>Psychiatric emergency<br>unit Jan-Mar |
| --- | --- | --- | --- | --- | --- | --- |
| Upper sec × 2016 | -0.276<br>[-0.688,0.137]<br>(0.191) | -3.696***<br>[-4.077,-3.315]<br>(0.000) | 0.483*<br>[0.077,0.890]<br>(0.020) | -0.318*<br>[-0.572,-0.064]<br>(0.014) | -2.239***<br>[-2.457,-2.022]<br>(0.000) | -0.094<br>[-0.351,0.163]<br>(0.474) |
| Upper sec × 2017 | 0.451*<br>[0.029,0.873]<br>(0.036) | -0.098<br>[-0.624,0.427]<br>(0.714) | 0.106<br>[-0.307,0.518]<br>(0.616) | 0.125<br>[-0.130,0.379]<br>(0.338) | -0.277<br>[-0.573,0.019]<br>(0.066) | -0.034<br>[-0.279,0.212]<br>(0.789) |
| Upper sec × 2018 | 0.109<br>[-0.311,0.529]<br>(0.612) | 0.314<br>[-0.220,0.847]<br>(0.249) | 0.191<br>[-0.221,0.604]<br>(0.363) | 0.197<br>[-0.061,0.454]<br>(0.134) | 0.150<br>[-0.160,0.459]<br>(0.343) | -0.019<br>[-0.270,0.232]<br>(0.881) |
| Upper sec × 2019 | ref | 0.502<br>[-0.036,1.039]<br>(0.067) | -0.266<br>[-0.689,0.157]<br>(0.218) | 0.000<br>[0.000,0.000]<br>(.) | 0.451**<br>[0.133,0.769]<br>(0.005) | 0.024<br>[-0.230,0.278]<br>(0.850) |
| Upper sec × 2020 | -0.895***<br>[-1.301,-0.488]<br>(0.000) | ref | ref | -0.379**<br>[-0.620,-0.137]<br>(0.002) | ref | ref |
| Upper sec × 2021 |  | -1.211***<br>[-1.743,-0.679]<br>(0.000) | -0.887***<br>[-1.291,-0.483]<br>(0.000) |  | -0.298*<br>[-0.594,-0.001]<br>(0.049) | -0.245*<br>[-0.481,-0.010]<br>(0.041) |
| Mean dep var upper sec 2019 | 4.92 | 8.45 | 4.89 | 2.24 | 3.28 | 2.20 |
| Effect in % | -18.19 | -14.33 | -18.14 | -16.92 | -9.08 | -11.16 |
| R2 | 0.002 | 0.004 | 0.002 | 0.001 | 0.002 | 0.001 |
| N | 3,276,398 | 3,985,925 | 3,985,925 | 3,276,398 | 3,985,925 | 3,985,925 |

Note: Results from linear regression model specified in main text. Models adjust for: sex, birth month, newly immigrated to Sweden, parental foreign background, income, education, income from social security systems, family size and if the biological parents live in the same household. Reference period in (1) and (4) are April-March 2019, in (2) and (5) July-December 2019 and in (3) and (6) January-March 2021.

